# Real-world effectiveness of Azvudine in hospitalized patients with COVID-19: a retrospective cohort study

**DOI:** 10.1101/2023.01.23.23284899

**Authors:** Minxue Shen, Chenggen Xiao, Yuming Sun, Daishi Li, Ping Wu, Liping Jin, Qingrong Wu, Yating Dian, Yu Meng, Furong Zeng, Xiang Chen, Guangtong Deng

## Abstract

Current guidelines prioritize the use of the Azvudine in coronavirus disease 2019 (COVID-19) patients. However, the clinical effectiveness of Azvudine in real-world studies was lacking, despite the clinical trials showed shorter time of nucleic acid negative conversion. To evaluate the clinical effectiveness following Azvudine treatment in hospitalized COVID-19 patients, we identified 1505 hospitalized COVID-19 patients during the study period, with a follow-up of up to 29 days. After exclusions and propensity score matching, we included 226 Azvudine recipients and 226 matched controls. The lower crude incidence rate of composite disease progression outcome (4.21 vs. 10.39 per 1000 person-days, *P*=0.041) and all-cause mortality (1.57 vs. 6.00 per 1000 person-days, *P*=0.027) were observed among Azvudine recipients compared with matched controls. The incidence rates of initiation of invasive mechanical ventilation were also statistically different between the groups according to the log-rank tests (*P*=0.020). Azvudine treatment was associated with significantly lower risks of composite disease progression outcome (hazard ratio [HR]: 0.43; 95% confidence interval [CI]: 0.18 to 0.99) and all-cause death (HR: 0.26; 95% CI: 0.07 to 0.94) compared with matched controls. Subgroup analyses indicated robustness of the point estimates of HRs (ranged from 0.14 to 0.84). Notably, male Azvudine recipients had a stronger effectiveness than female recipients with respect to both composite outcome and all-cause death. These findings suggest that Azvudine treatment showed substantial clinical benefits in hospitalized COVID-19 patients, and should be considered for use in this population of patients.

## INTRODUCTION

Coronavirus disease 2019 (COVID-19) remains a major threat to global health, especially in China, as COVID-19 prevention and control measures change^1^. Numerous drugs have been repurposed or developed to treat COVID-19 patients, including Azvudine, the first Chinese oral anti-COVID-19 drug^2,3^. Azvudine has been reported to shorten the time of nucleic acid negative conversion in the mild and common COVID-19^4^. Besides, Azvudine cured all patients with common and severe COVID-19 in a randomized, single-arm clinical trial^5^. An unpublished phase 3 multicenter randomized clinical study further suggested that Azvudine significantly shorten the symptom improvement time and increase the proportion of COVID-19 patients with improved clinical symptoms^6^. Thus, Azvudine has been approved and recommended to treat COVID-19 patients in the Diagnosis and Treatment Program for Novel Coronavirus Pneumonia^7,8^.

Although current guidelines prioritize the use of the Azvudine in common COVID-19 patients, several concerns and research gaps remain, such as whether Azvudine reduces the adverse clinical outcomes in hospitalized COVID-19 patients, the need for more clinical data on the real-world effectiveness. In particular, the previous clinical trials mainly focus on the time of nucleic acid negative conversion and symptom improvement, and the effectiveness of Azvudine in disease progression and mortality has been questioned due to the delays in disclosure of results from multi-center randomized clinical trials with large sample sizes^4-6^. To close this data gap, real-world evidence of the effectiveness of Azvudine in COVID-19 patients is urgently needed.

In this retrospective cohort study, we aimed to evaluate the clinical effectiveness following the use of Azvudine in hospitalized COVID-19 patients in Xiangya hospital, the largest hospital located in the south-central region of China. Although Azvudine is now indicated for common COVID-19 patients, the current analysis focuses on their effectiveness in hospitalized COVID-19 patients who do not initially require any oxygen therapy on admission.

## RESULTS

We consecutively collected data on 1505 patients with a confirmed diagnosis of SARS-CoV-2 infection who were admitted to Xiangya hospital, with a follow-up of up to 29 days. A total of 226 Azvudine recipients and 674 controls who had no requirement for oxygen therapy at baseline were eligible for inclusion (Fig. 1). Baseline characteristics of the Azvudine and control groups before and after 1:1 propensity-score matching are presented in Table 1. After matching, we included 226 Azvudine recipients and 226 matched controls, and the baseline characteristics of patients were balanced between the two groups, with standard mean differences (SMDs) lower than 0.1 (Supplementary Fig. 1). As a result of shortage in medical resource and drug supply during this pandemic, the mean duration from symptom onset to hospitalization was 8.2 days, and only 12% patients received Azvudine within 5 days of symptom onset. Frequencies of symptoms (Supplementary Fig. 2) and laboratory parameters (Supplementary Table 1) were comparable between the groups in general.

**Figure 1.**
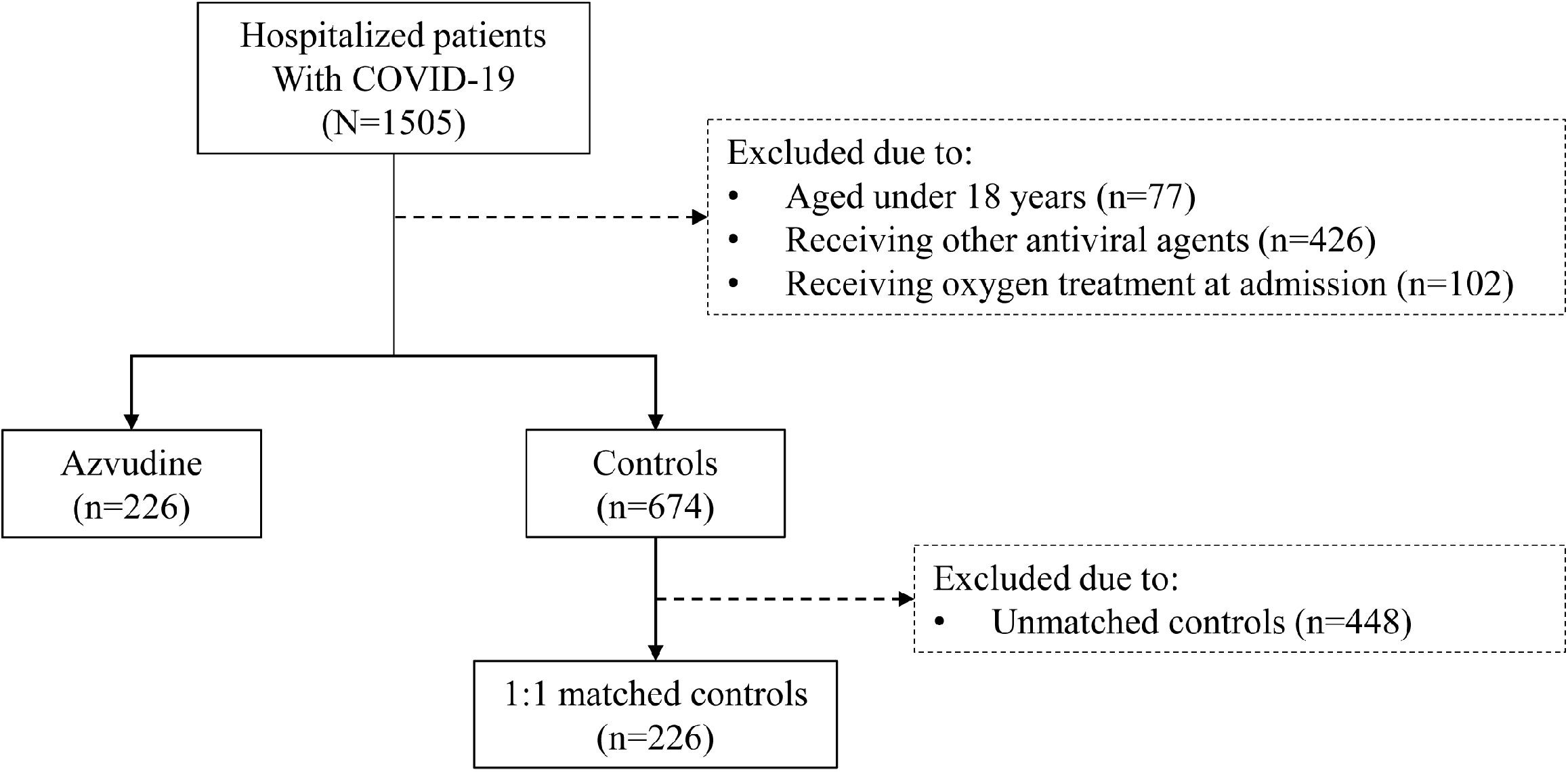
Identification of Azvudine recipients and their matched controls among COVID-19 hospitalized patients during the study period.

**Table 1.**
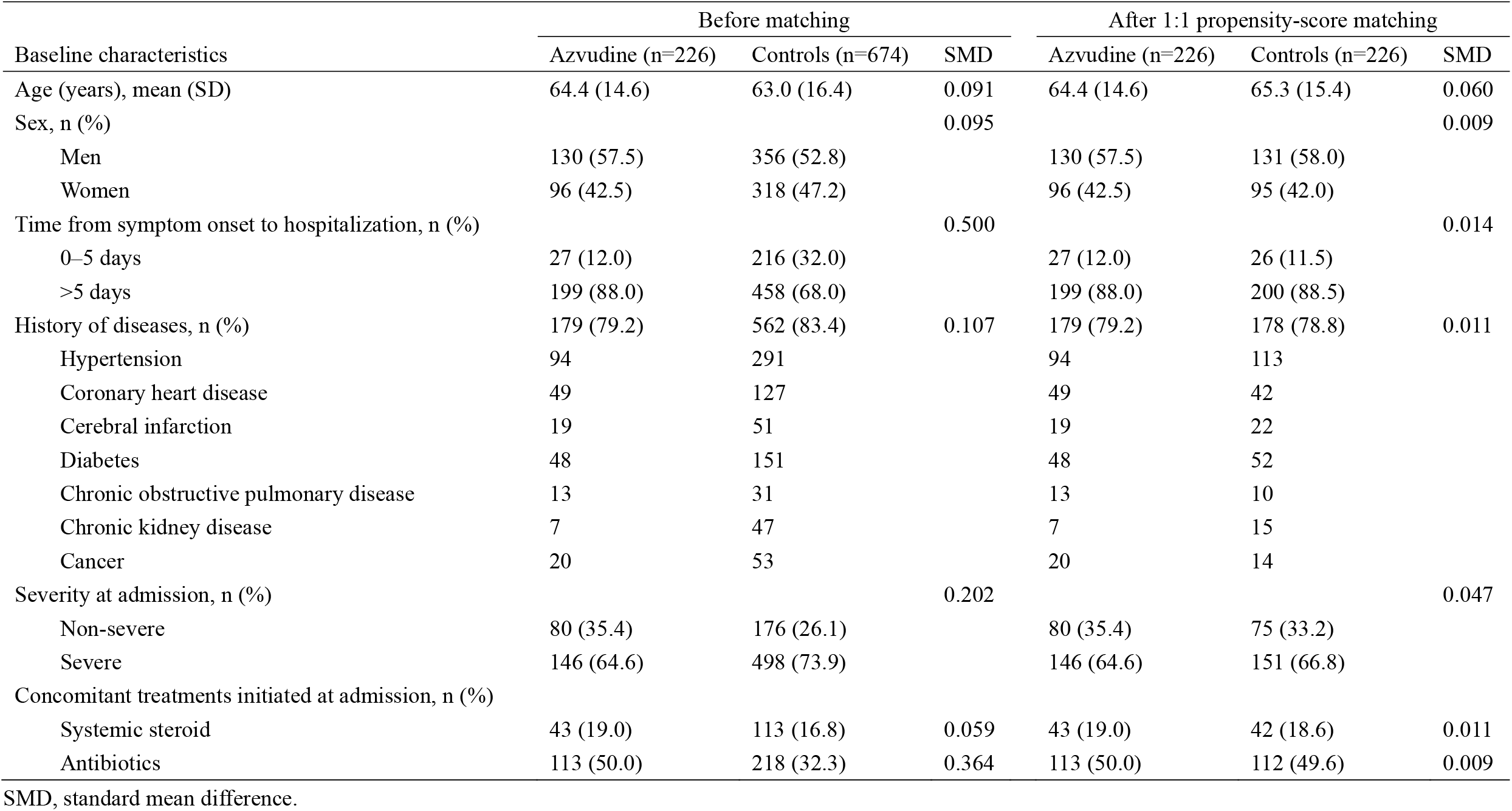
Baseline characteristics of the participants.

The crude incidence rates of composite disease progression outcome were 4.21 per 1000 person-days in patients treated with Azvudine versus 10.39 per 1000 person-days in the control group (*P*=0.041). The crude all-cause mortality rate was 1.57 per 1000 person-days among Azvudine recipients and 6.00 per 1000 person-days among the controls (*P*=0.027). The incidence rates of initiation of invasive mechanical ventilation were also statistically different between the groups according to the log-rank tests (*P*=0.020) (Table 2). Azvudine treatment was associated with significantly lower risks of composite outcome (hazard ratio [HR]: 0.43; 95% confidence interval [CI]: 0.18 to 0.99) and all-cause death (HR: 0.26; 95% CI: 0.07 to 0.94) compared with controls (Fig. 2, 3). The HR for invasive mechanical ventilation was not estimated because no outcome was observed among Azvudine recipients. In contrast, treatment with Azvudine was not significantly associated with intensive care unit admission and high-flow oxygen therapy.

**Table 2.** Composite and individual outcomes in Azvudine group versus matched controls.

| Outcome | Azvudine (n=226) |  | Matched controls (n=226) |  | *P*<sup>a</sup> |
| --- | --- | --- | --- | --- | --- |
|  | n (%) | Rate per 1000 person-days | n (%) | Rate per 1000 person-days |  |
| Composite outcome, n (%) | 8 (3.54) | 4.21 | 17 (7.52) | 10.39 | 0.041 |
| All-cause death, n (%) | 3 (1.33) | 1.57 | 10 (4.42) | 6.00 | 0.027 |
| Intensive care unit admission, n (%) | 1 (0.44) | 0.51 | 4 (1.77) | 2.32 | 0.149 |
| Initiation of invasive mechanical ventilation, n (%) | 0 | ... | 5 (2.21) | 2.90 | 0.020 |
| Need for high-flow oxygen therapy, n (%) | 6 (2.65) | 3.04 | 7 (3.10) | 4.06 | 0.641 |

<sup>a</sup> *P* value for log-rank test.

**Figure 2.**
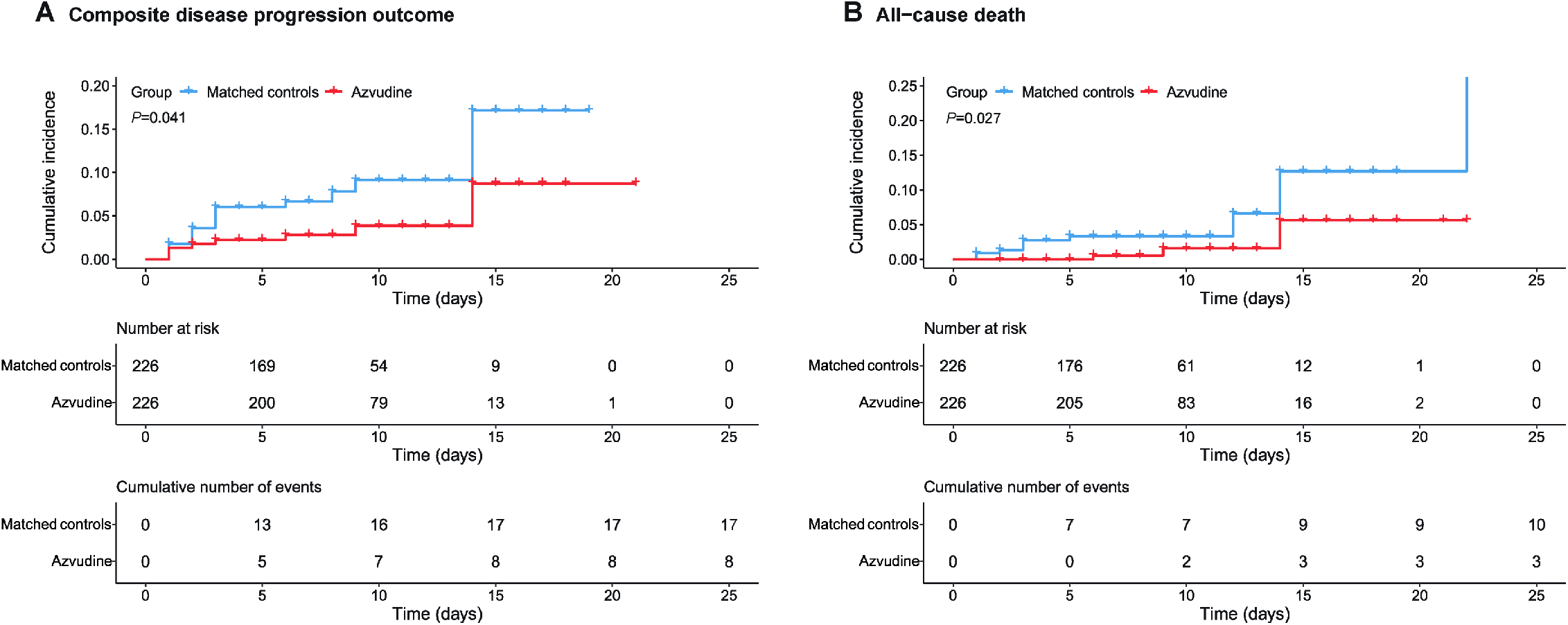
Cumulative incidence of composite disease progression outcome (A) and all-cause death (B) for Azvudine recipients versus matched controls.

**Figure 3.**
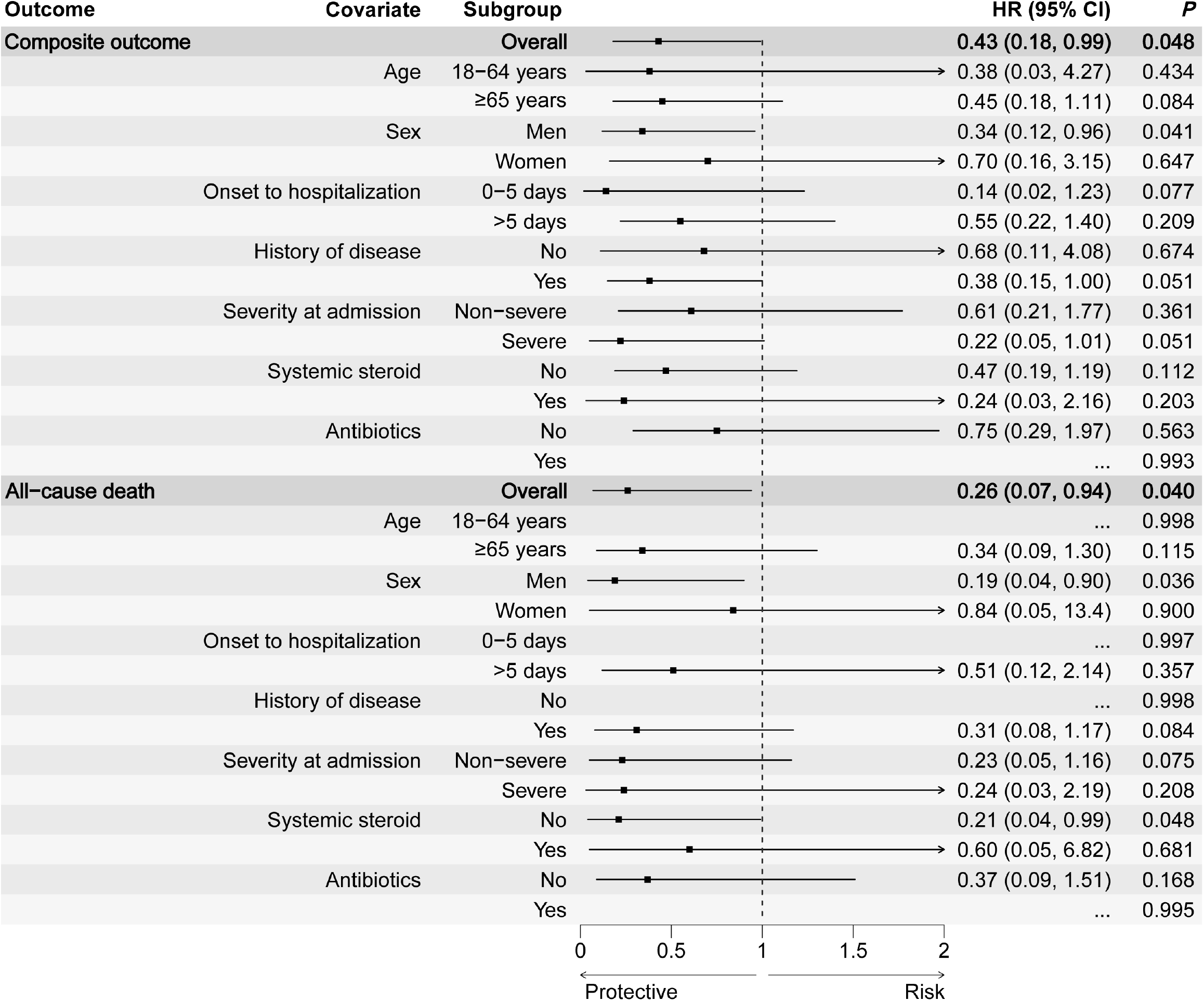
The effectiveness of Azvudine in reducing the risk of composite disease progression outcome and all-cause death by subgroups of selected baseline characteristics.

Results of subgroup analyses indicated robustness of the point estimates of HRs (ranged from 0.14 to 0.84); however, the 95% CIs were broad due to the sample size. It is noteworthy that male recipients of Azvudine had a stronger effectiveness than female recipients with respect to both composite outcome and all-cause death (Fig. 3).

## DISCUSSION

At the beginning of the change in COVID-19 prevention and control measures in December 2022^1^, two oral antiviral drugs were approved and recommended to treat the COVID-19 patients in the Diagnosis and Treatment Program for Novel Coronavirus Pneumonia including nirmatrelvir-ritonavir and Azvudine^7-9^. Nirmatrelvir-ritonavir has been reported to reduce severe COVID-19 and mortality in high-risk patients in clinical trials and real-world studies^10-12^. Nevertheless, the clinical effectiveness of Azvudine in real-world studies is lacking, despite the clinical trials showed shorter time of nucleic acid negative conversion.

In the retrospective cohort of hospitalized COVID-19 patients who do not initially require any oxygen therapy on admission, Azvudine administration was associated with significantly lower risk of composite disease progression outcome and all-cause death. To our knowledge, this is the first real-world study exploring the inpatient use of the oral antiviral drug during the pandemic wave in China. Although the 95% CIs are broad, the direction of the results is in line with the results of previous clinical trials that provide evidence upon which the National Medical Products Administration based its decision to grant Azvudine to treat COVID-19.

In the interim analysis of an unpublished phase 3 multicenter randomized clinical study, 40.43% COVID-19 patients following Azvudine treatment had improved clinical symptoms, compared with 10.87% control patients^6^. In our study, the crude incidence rate of all-cause death was 1.33% among Azvudine recipients and 4.42% among the controls, and the rates of composite outcome of the two groups were 3.54% and 7.52%, respectively. The consistent results further reflect the clinical effectiveness of Azvudine in the real-world clinical practice and provide real-world evidence supporting their use in COVID-19 patients.

Azvudine is a 4’-modified nucleoside whose active triphosphate was embedded into viral RNA during SARS-CoV-2 RNA synthesis, ultimately terminating viral RNA replication^13,14^. Interestingly, we found that male recipients of Azvudine had a stronger effectiveness than female recipients with respect to both composite outcome and all-cause death. The findings were consistent with previous results in vitro from molnupiravir, another nucleoside-based RdRp inhibitor for COVID-19 treatment. Lieber et al. found that lung virus titer reductions in molnupiravir-treated males were highly significant compared to vehicle-treated males, but not in molnupiravir-treated females^15^. However, the potential mechanism by which male COVID-19 patients benefit better from nucleoside-based RdRP inhibitors requires further investigation. A strength of the current study is that we used the medical records of hospitalized patients, thus close monitoring, and the clinical outcomes and procedures were therefore systematically documented and analyzed. Medication adherence could also be guaranteed in inpatients compared outpatients. However, this study has other limitations. First, we cannot exclude the possibility of selection bias or confounding by indication in the retrospective cohort study, despite the data were consecutively collected and adjusted for a large number of confounders associated with high risk for severe COVID-19. Second, we did not evaluate the time of nucleic acid negative conversion, because the Ct value was no longer being used as a discharge criterion and not checked regularly during the special period. Third, this was a single-center study with a small sample size and marginal significance as the primary outcome. Considering the surge of the omicron variant in China, we adopted the method of interim analysis to evaluate the drug effect under the premise of minimum sample size, so that the study results can be rapidly applied to clinical practice and improve the prognosis of large-scale patients in China. Fourth, the generalizability of our findings could be undermined by the inpatient setting of our cohort, and some of our subgroup analyses were more likely to be underpowered due to the small sample size. Finally, the evaluation of adverse events and safety data reports was beyond the scope of this study. It should be noted that Azvudine is not recommended in several clinical contraindications associated with drug-drug interactions and those with severe renal or liver diseases. Therefore, clinicians should carefully assess the severity of the patient’s diseases, review the patient’s concomitant medications, and evaluate potential drug-drug interactions. Future studies will be needed to assess the short- and long-term safety of Azvudine in real-world settings.

In conclusion, our findings suggest that Azvudine treatment is associated with significantly lower risks of composite disease progression outcome and all-cause death in real-world clinical practice, especially in male COVID-19 patients.

## METHODS

### Study design

We conducted a single-center, retrospective cohort study of hospitalized adult patients with positive RT-PCR for SARS-CoV-2 infection, who were given standard treatment or Azvudine plus standard treatment in Xiangya Hospital, during the period from Dec 5, 2022 to Jan 13, 2023. We excluded patients who were younger than 18 years; those receiving antiviral agents other than Azvudine; and those with oxygen support or mechanical ventilation on the date of admission. This study was approved by the institutional review board of Xiangya Hospital, Central South University (202002024). Individual patient-informed consent was not required for this retrospective cohort study using anonymized data.

### Data source

Electronic health records of patients with COVID-19 were retrieved from the inpatient system of Xiangya Hospital. The records include demographic characteristics, data on admissions, diagnoses, prescription and drug dispensing records, procedures, and laboratory tests, and date of discharge or death. The data were consecutively collected until a significant result was obtained in interim analyses when the total sample size reached 500, 1000, and 1500, respectively.

### Treatment exposure

Hospitalized patients with COVID-19 who received Azvudine treatment during the observation period were defined as having treatment exposure. We defined the treatment exposure period as within the first 2 days of admission to mitigate potential immortal time bias between treatment initiation and admission. Controls were selected from the hospitalized patients with COVID-19 who did not receive Azvudine or other antiviral agents during the observation period, using propensity-score matching in a ratio of 1:1.

### Outcomes

The primary outcome was a composite outcome of disease progression including all-cause death, intensive care unit admission, initiation of invasive mechanical ventilation, and need for high-flow oxygen therapy, and the secondary outcome was each of these individual disease progression outcomes. Patients were observed from the admission date until the occurrence of outcome events, the date of discharge, or the date of death, whichever came first.

### Baseline covariates

Baseline covariates of patients included age, sex, time from symptom onset to hospitalization, history of diseases, severity of COVID-19 on admission (severe cases were defined as having respiratory rate ≥30, or oxygen saturation ≤93%, or PaO2/FiO2 ≤300 mmHg, or lung infiltrates >50%), concomitant treatments initiated at admission (systemic steroid and antibiotics). Symptoms and laboratory parameters on admission were also collected and described.

### Statistical analysis

We used propensity-score models conditional on the aforementioned baseline covariates, and the probability of receiving Azvudine was estimated in an approach of calliper matching without replacement, with a calliper width of 0.2. We used the SMDs to assess the balance of each baseline covariate between the groups before and after propensity-score matching, with an SMD greater than 0.1 indicating covariate imbalance^16,17^. HRs with 95% CIs for each outcome between the groups were estimated using Cox regression models. Subgroup analyses were conducted in each stratum of the aforementioned baseline covariates to evaluate the robustness of the estimates. All statistical analyses were done with R version 4.2.1. The level of significance was two-tailed 0.05 for statistical tests.

## Data Availability

All data produced in the present study are available upon reasonable request to the authors

## ACKNOWLEDGEMENTS

We thank all the hospital staff members for their efforts in collecting the information that used in this study; thank the patients who participated in this study, their families, and the medical, nursing, and research staff at the study centers.

## CONTRIBUTIONS

Conception and design: Guangtong, Xiang Chen, Furong Zeng, and Minxue Shen. Acquisition of data: Furong Zeng, Chenggen Xiao, Yuming Sun, Daishi Li, Ping Wu. Interpretation of data, statistical analysis and manuscript writing: Guangtong Deng, Furong Zeng, and Minxue Shen. Revision of manuscript and administrative, technical, or material support: Guangtong Deng, Furong Zeng, Xiang Chen, Minxue Shen, Yu Meng, Yating Dian.

