## Supplementary materials for "Real-world effectiveness of Azvudine in hospitalized patients with COVID-19: a retrospective cohort study"

**This PDF file includes:**

Figures. S1 to S2
Tables S1

**Figure. S1.**

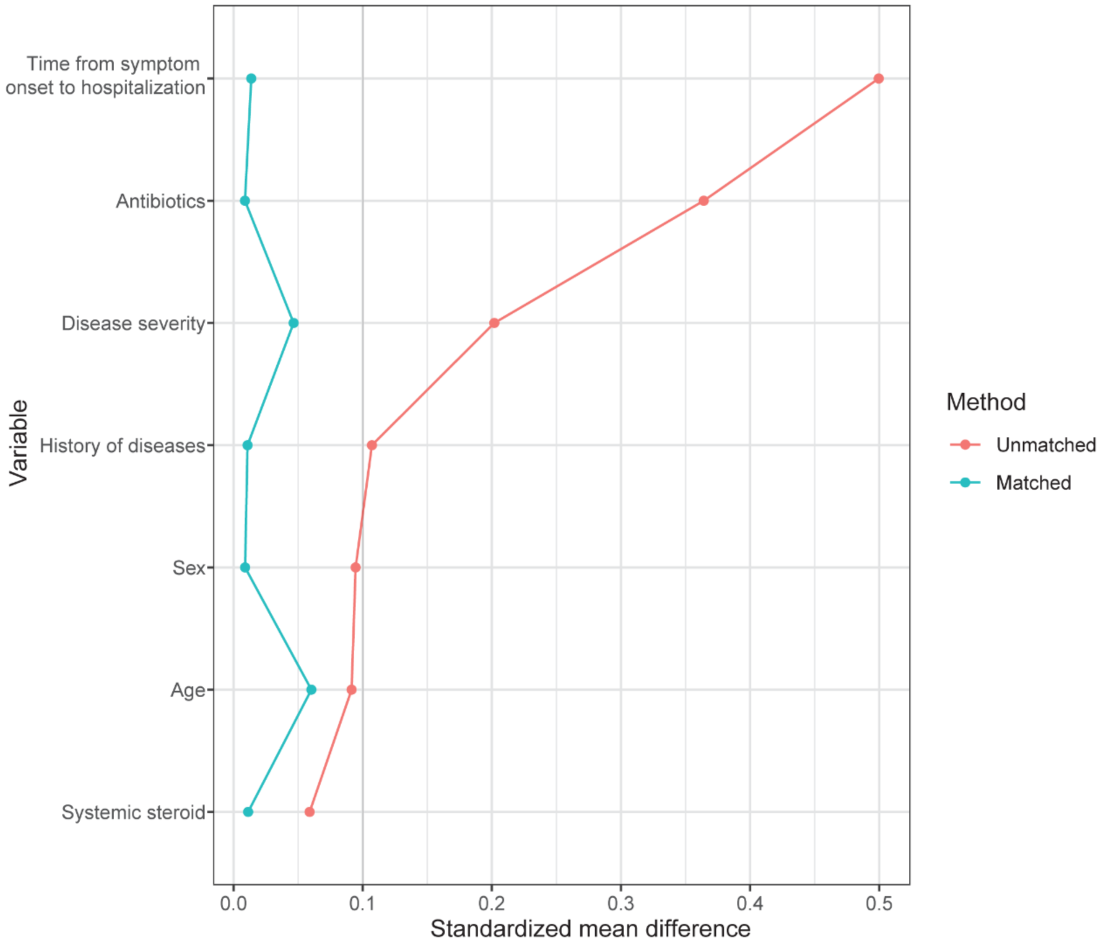


Standard mean differences between the groups before and after 1:1 propensity score-matching.

**Figure. S2.**

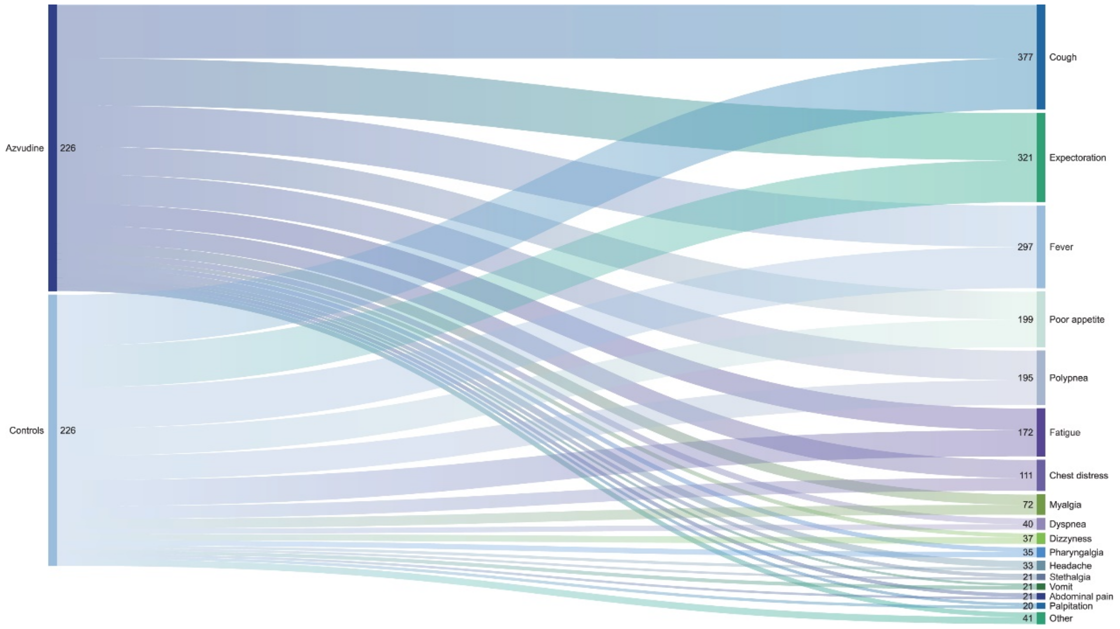


Frequency of symptoms between Azvudine recipients and matched controls.

**Table S1. Laboratory parameters between the two groups.**

| Category | Indicator | Azvudine | | Control | |
|---|---|---|---|---|---|
| | | Mean | SD | Mean | SD |
| Arterial blood gas analysis | $PaO_2$ | 108.4 | 46.7 | 108.0 | 53.7 |
| | $PaCO_2$ | 36.0 | 7.0 | 36.4 | 7.6 |
| | $PaO_2/FiO_2$ | 388.4 | 173.6 | 379.5 | 181.3 |
| Blood cell count | Red blood cell | 4.1 | 0.6 | 6.9 | 34.2 |
| | Hemoglobin | 128.4 | 76.0 | 118.1 | 19.3 |
| | White blood cell | 6.2 | 2.9 | 6.3 | 2.9 |
| | Blood platelet | 228.8 | 289.7 | 233.4 | 104.2 |
| | Lymphocyte | 1.1 | 0.6 | 1.2 | 1.2 |
| | Lymphocyte (%) | 20.8 | 10.9 | 20.6 | 11.5 |
| | Neutrophil | 4.6 | 4.1 | 5.1 | 6.8 |
| | Neutrophil (%) | 68.3 | 12.6 | 68.2 | 14.3 |
| | Eosinophil | 0.1 | 0.2 | 0.1 | 0.2 |
| | Basophil | 0.0 | 0.1 | 0.0 | 0.2 |
| Liver, renal, and cardiac function | Total bilirubin | 10.8 | 4.5 | 11.8 | 22.4 |
| | Direct bilirubin | 3.9 | 1.7 | 4.8 | 14.3 |
| | Albumin | 35.3 | 4.1 | 34.7 | 4.5 |
| | Globulin | 28.3 | 4.5 | 28.7 | 4.9 |
| | ALT | 31.8 | 29.8 | 32.3 | 27.2 |
| | AST | 30.6 | 18.8 | 34.8 | 30.5 |
| | Blood creatinine | 89.2 | 114.7 | 113.2 | 156.2 |
| | Blood urea nitrogen | 6.4 | 5.2 | 7.3 | 5.7 |
| | $K^+$ | 4.0 | 0.8 | 4.0 | 0.5 |

| | | | | | |
|---|---|---|---|---|---|
| | $Ca^{2+}$ | 2.2 | 1.2 | 2.1 | 0.1 |
| | $Na^{+}$ | 145.6 | 86.2 | 144.9 | 84.8 |
| | $Cl^{-}$ | 103.6 | 6.7 | 152.8 | 668.1 |
| | $Mg^{2+}$ | 1.4 | 6.7 | 0.8 | 0.1 |
| | Lactic dehydrogenase | 228.5 | 81.0 | 220.9 | 96.1 |
| | Creatine kinase | 102.9 | 180.7 | 141.7 | 449.7 |
| | creatine kinase MB | 11.9 | 10.3 | 10.4 | 5.8 |
| | Myohemoglobin | 80.6 | 100.9 | 132.6 | 432.1 |
| | Troponin | 3.4 | 40.2 | 2.1 | 22.6 |
| | Brain natriuretic peptide | 609.0 | 1556.3 | 1431.7 | 4457.2 |
| Coagulation | D-Dimer | 0.3 | 0.9 | 0.5 | 1.7 |
| | PT | 12.1 | 2.6 | 12.2 | 2.6 |
| | INR | 1.0 | 0.3 | 1.0 | 0.2 |
| | Fibrinogen | 4.4 | 1.5 | 4.6 | 1.8 |
| | APTT | 27.5 | 4.2 | 28.5 | 6.8 |
| | TT | 16.6 | 4.9 | 16.4 | 1.9 |
| Cytokines | TNF-α | 10.0 | 6.3 | 8.4 | 7.4 |
| | Interferon-γ | 2.5 | 0.0 | 5.4 | 2.3 |
| | IL-2 | 1.9 | 1.0 | 1.9 | 1.1 |
| | IL-4 | 2.5 | 0.0 | 2.5 | 0.0 |
| | IL-6 | 19.6 | 32.9 | 34.2 | 133.6 |
| | IL-10 | 6.3 | 7.9 | 5.9 | 14.3 |
| | IL-17A | 1.7 | 0.0 | 1.4 | 0.5 |
| Other | $HbA_{1c}$ | 6.7 | 1.3 | 6.5 | 1.6 |
| | Lactic acid | 1.6 | 0.7 | 1.9 | 1.2 |
| | Procalcitonin | 0.3 | 1.8 | 1.7 | 14.9 |

| | | | | | |
|---|---|---|---|---|---|
| | C-reactive protein | 42.2 | 51.9 | 38.0 | 47.7 |
| | Erythrocyte sedimentation rate | 54.1 | 26.9 | 58.2 | 29.3 |
| | Ferritin | 791.2 | 544.0 | 575.7 | 487.3 |

SD, standard deviation.
